# Low uptake of COVID-19 prevention behaviours and high socioeconomic impact of lockdown measures in South Asia: evidence from a large-scale multi-country surveillance programme

**DOI:** 10.1101/2020.11.12.20229898

**Authors:** Dian Kusuma, Rajendra Pradeepa, Khadija I Khawaja, Mehedi Hasan, Samreen Siddiqui, Sara Mahmood, Syed Mohsin Ali Shah, Chamini K De Silva, Laksara de Silva, Manoja Gamage, Menka Loomba, Vindya P Rajakaruna, Abu AM Hanif, Rajan Babu Kamalesh, Balachandran Kumarendran, Marie Loh, Archa Misra, Asma Tassawar, Akansha Tyagi, Swati Waghdhare, Saira Burney, Sajjad Ahmad, Viswanathan Mohan, Malabika Sarker, Ian Y Goon, Anuradhani Kasturiratne, Jaspal S Kooner, Prasad Katulanda, Sujeet Jha, Ranjit Mohan Anjana, Malay K Mridha, Franco Sassi, John C Chambers, on behalf of the NIHR Global Health Research Unit for Diabetes and Cardiovascular Disease in South Asia

**Affiliations:** Centre for Health Economics & Policy Innovation, Imperial College Business School, UK; Madras Diabetes Research Foundation, Chennai, India; Services Institute of Medical Sciences, Lahore, Pakistan; BRAC James P Grant School of Public Health, BRAC University, Dhaka, Bangladesh; Max Healthcare, New Delhi, India; Punjab Institute of Cardiology, Lahore, Pakistan; Faculty of Medicine, University of Kelaniya, Ragama, Sri Lanka; Faculty of Medicine, University of Colombo, Colombo, Sri Lanka; Faculty of Medicine, University of Jaffna, Jaffna, Sri Lanka; Lee Kong Chian School of Medicine, Nanyang Technological University, Singapore, Singapore; School of Public Health, Imperial College London, London, UK; National Heart and Lung Institute, Imperial College London, London, UK

**Author notes:** **Correspondence to** John C Chambers, Department of Epidemiology and Biostatistics, School of Public Health, Imperial College London, London W2 1PG, UK.

## Abstract

**Background:** South Asia has become a major epicentre of the COVID-19 pandemic. Understanding South Asians’ awareness, attitudes and experiences of early measures for the prevention of COVID-19 is key to improving the effectiveness and mitigating the social and economic impacts of pandemic responses at a critical time for the Region.

**Methods:** We assessed the knowledge, behaviours, health and socio-economic circumstances of 29,809 adult men and women, at 93 locations across four South Asian countries. Data were collected during the national lockdowns implemented from March to July 2020, and compared with data collected prior to the pandemic as part of an ongoing prospective surveillance initiative.

**Results:** Participants were 61% female, mean age 45.1 years. Almost half had one or more chronic disease, including diabetes (16%), hypertension (23%) or obesity (16%). Knowledge of the primary COVID-19 symptoms and transmission routes was high, but access to hygiene and personal protection resources was low (running water 63%, hand sanitisers 53%, paper tissues 48%). Key preventive measures were not widely adopted. Knowledge, access to, and uptake of COVID-19 prevention measures were low amongst people from disadvantaged socio-economic groups. Fifteen percent of people receiving treatment for chronic diseases reported loss of access to long-term medications; 40% reported symptoms suggestive of anxiety or depression. The prevalence of unemployment rose from 9.3% to 39.4% (P<0.001), and household income fell by 52% (P<0.001) during the lockdown. Younger people and those from less affluent socio-economic groups were most severely impacted. Sedentary time increased by 32% and inadequate fruit and vegetable intake increased by 10% (P<0.001 for both), while tobacco and alcohol consumption dropped by 41% and 80%, respectively (P<0.001), during the lockdown.

**Conclusions:** Our results identified important knowledge, access and uptake barriers to the prevention of COVID-19 in South Asia, and demonstrated major adverse impacts of the pandemic on chronic disease treatment, mental health, health-related behaviours, employment and household finances. We found important sociodemographic differences for impact, suggesting a widening of existing inequalities. Our findings underscore the need for immediate large-scale action to close gaps in knowledge and access to essential resources for prevention, along with measures to safeguard economic production and mitigate socio-economic impacts on the young and the poor.

## Background

South-Asia is the most densely populated region of the world (1.9 billion people, 25% of global population), with more than 98% of South Asians living in Bangladesh, India, Pakistan or Sri Lanka. In common with residents of many other lower-middle income countries (LMICs), the people of South Asia face multiple challenges, including high rates of both communicable and non-communicable disease, fragile health and education systems, food and financial insecurity, and limited formal economic or social support.[1,2] Together, these characteristics are anticipated to make South Asian countries more vulnerable to major health and societal challenges such as COVID-19.

The first case of COVID-19 in South Asia was identified in January 2020, shortly before COVID-19 was declared a pandemic by the WHO.[3] In response, Bangladesh, India, Pakistan and Sri Lanka implemented a range of highly restrictive national control measures to reduce the spread of COVID-19. This included closures of schools and non-essential workplaces, public transport bans, education campaigns for individual level behavioural interventions, isolation of symptomatic individuals, and contact tracing.[4,5] However, since then more than 7.4 million cases have been detected, with over 116,500 deaths (7th October 2020, https://www.worldometers.info/coronavirus/). The rate of new infections is still rapidly accelerating in India, which now has the second highest numbers of COVID-19 cases globally, indicating that the control measures may have been less effective than in other settings. Furthermore, there is increasing evidence that national lockdowns may have adverse effects for physical and mental health, children’s education, behaviours relevant to chronic disease, as well as severe social and financial consequences.[6–10] There is an urgent need to understand how control measures can be further strengthened in South Asia, both to reduce the high rates of ongoing transmission, and to mitigate their unintended adverse consequences.

We interviewed 29,809 people participating in a long-term South Asian health surveillance study while restrictive national control measures were in place. We measured their knowledge of COVID-19, adoption of preventive practices, and impact of COVID-19 on their physical and mental health, health-related behaviours, non-communicable disease care, social circumstances, and economic well-being. The ultimate goal of the study was to inform the design and implementation of further COVID-19 prevention and control programs in South Asia.

## Methods

We used our network of health surveillance sites to evaluate the impact of COVID-19 on people living in five study regions: Bangladesh, South India, North India, Pakistan, and Sri Lanka. The surveillance sites include 52,813 South Asian men and women aged 18 years and above with comprehensive information on baseline health collected immediately prior to the COVID-19 pandemic (November 2018 to March 2020). We supplemented these baseline data with a telephone interview focussed on COVID-19, which was completed by 29,809 of the surveillance study participants, during the national lockdowns implemented between March and July 2020. The study was approved by IRBs in each country, and consent was obtained from all participants for each round of data collection.

### Study settings and recruitment

Recruitment to our health surveillance study took place at 93 surveillance sites (range 2 to 33 per study region, 74% urban, **Supplementary Table 1**). Governmental census data and available household listings were used, together with house-house visits by research teams and local primary care workers, to identify (enumerate) the resident population. Resident adults (age 18 years and above) were invited to take part; exclusion criteria included current pregnancy, or serious illness expected to reduce life expectancy to less than 12 months. We worked closely with community senior members (e.g. teachers, employers, religious leaders) to support and facilitate engagement in the study. Explanations of the project’s purpose were provided in writing and using videos, in relevant South Asian languages, supported by bilingual translators. Recruitment started in November 2018, and by March 2020 we had completed evaluation of 52,813 people (Bangladesh: 13,955; North India: 9,469; South India: 8,621; Pakistan: 5,833; Sri Lanka: 14,935).

### Baseline evaluation

An interviewer-administered health and lifestyle questionnaire was used to collect information on behavioural risk factors (smoking, alcohol use, physical activity and consumption of fruits/vegetables), personal and family medical history, medications, and socio-economic status. Physical measurements included: a) Anthropometry (height, weight, waist and hip circumference and bio-impedance for body fat composition); b) Blood pressure by Omron digital device; c) Cardiac evaluation by 12 lead ECG to identify arrhythmia, left ventricular hypertrophy and previous myocardial infarction; d) Retinal photography for assessment of retinal disease, including hypertensive and diabetic retinopathy; and e). Respiratory evaluation by spirometry to assess for smoking/environment-related lung injury. Fasting glucose, and cholesterol were measured by point of care tests. Aliquots of blood and urine are stored for future molecular analyses. Equipment, protocols and training was standardised across sites.

### Follow-up COVID-19 questionnaire

As part of an integrated effort, co-ordinated by the Wellcome Trust, we developed a questionnaire aimed at assessing (i) prevalence, knowledge and uptake of behaviours relevant to COVID-19; and, (ii) impact of COVID-19 on access to healthcare, behaviours linked to non-communicable disease risk, social interactions and financial circumstances.[11] The questionnaire is available in full on our study website (www.ghru-southasia.org). We implemented the questionnaire by telephone during the national lockdown period, using a bespoke data capture tool based on KoBoToolbox, an open source data collection software. The study team attempted to contact all 52,813 participants of our surveillance study. Training for questionnaire administration, and quality control of data collection were co-ordinated centrally.

### Statistical analysis

Descriptive statistics are presented as mean (SD) or as % for continuous and categorical variables, respectively. Participants were clustered geographically in surveillance sites. To account for heterogeneity in outcomes between sites due to unobserved contextual factors, we used a multilevel modelling regression approach with random effects (intercept) at the study site level to quantify the relationships between outcomes and potential predictors such as age, gender, education, prior income, and prior chronic condition. For analyses where the baseline (pre-pandemic) and follow-up (during the pandemic) data (e.g. behaviours linked with chronic disease risk, social and financial circumstances) were available, we examined the changes between the two time points. We conducted all analyses in STATA 15.

We analysed three groups of dependent variables: (i) COVID-19 burden, knowledge and preventive behaviours of COVID-19, (ii) Impact of COVID-19 on chronic disease and behaviours linked with chronic disease risk, and (iii) Impact of COVID-19 on social and financial circumstances. The prevalence of COVID-19 was estimated from reported symptoms, such as fever and cough; knowledge was tested for disease symptoms and transmission mechanism; and preventive behaviours included washing hands, wearing masks, and following social distancing. Impact on chronic disease included running out of routine medications, symptoms of psychosocial distress (e.g. anxiety and depression), and risk factors for chronic conditions (e.g. alcohol drinking, tobacco use, physical in activity, and unhealthy diet). Finally, impact on social and financial circumstances included household income, employment, and working hours.

## Results

### Study population

The baseline characteristics of all 52,813 participants in our surveillance study are summarised in **Supplementary Table 2**. Of these, 29,809 people successfully completed the COVID-19 questionnaire (**Table 1**, response rate 57%), with little evidence for responder bias (**Supplementary Figure 1**). Participants were 61% female, mean age 45 years, and living in Bangladesh (8,807), North India (6,152), South India (3,834), Pakistan (2,534) or Sri Lanka (8,382). Almost half of participants had at least one chronic disease, including diabetes (16%), hypertension (23%) or obesity (16%). Participants from South India and Sri Lanka were older, and had higher prevalence of diabetes, compared to other settings (P<0.001). Cigarette smoking was highest, whilst education and indicators of socio-economic status were lowest, in Bangladesh and Pakistan (P<0.001). The ‘lockdown’ control measures for each country during the survey period included major restrictions on movement outside the house, closure of schools and non-essential workplaces, and cancellation of public and religious events (**Supplementary Table 3**).

**Table 1.** Characteristics of the 29,809 study participants prior to COVID-19 pandemic

|  | Total |  | Bangladesh |  | North India |  | South India |  | Pakistan |  | Sri Lanka |  |
| --- | --- | --- | --- | --- | --- | --- | --- | --- | --- | --- | --- | --- |
|  | n/mean | %/SD | n/mean | %/SD | n/mean | %/SD | n/mean | %/SD | n/mean | %/SD | n/mean | %/SD |
| (a) Basic characteristics |  |  |  |  |  |  |  |  |  |  |  |  |
| Female (n, %) | 18111 | 60.8% | 4622 | 52.5% | 3745 | 60.9% | 2330 | 59.2% | 1644 | 64.9% | 5770 | 68.8% |
| Age (year, SD) | 45.1 | 14.1 | 42.8 | 13.6 | 41.6 | 13.6 | 48.9 | 12.9 | 42.6 | 12.6 | 49.1 | 14.5 |
| Household income (USD, SD) | 240.0 | 203.9 | 219.2 | 188.7 | 307.9 | 247.9 | 234.0 | 179.4 | 176.6 | 184.7 | 235.3 | 187.7 |
| Education (year, SD) | 7.3 | 4.7 | 5.7 | 4.7 | 7.1 | 4.6 | 7.1 | 3.9 | 3.7 | 4.9 | 10.4 | 2.8 |
| Employment |  |  |  |  |  |  |  |  |  |  |  |  |
| Government employee (n, %) | 1064 | 3.6% | 156 | 1.8% | 83 | 1.4% | 95 | 2.4% | 213 | 8.5% | 517 | 6.2% |
| Non-government employee (n, %) | 4792 | 16.3% | 606 | 6.9% | 1724 | 29.0% | 1024 | 26.1% | 163 | 6.5% | 1275 | 15.4% |
| Self-employed (n, %) | 7624 | 25.9% | 3455 | 39.4% | 848 | 14.3% | 1300 | 33.2% | 344 | 13.7% | 1677 | 20.2% |
| Non-paid (n, %) | 60 | 0.2% | 3 | 0.03% | 20 | 0.3% | 17 | 0.4% | 6 | 0.2% | 14 | 0.2% |
| Student (n, %) | 729 | 2.5% | 157 | 1.8% | 274 | 4.6% | 23 | 0.6% | 61 | 2.4% | 214 | 2.6% |
| Homemaker (n, %) | 10953 | 37.2% | 3983 | 45.4% | 2551 | 42.9% | 1164 | 29.7% | 1525 | 60.9% | 1730 | 20.8% |
| Retired (n, %) | 1116 | 3.8% | 179 | 2.0% | 147 | 2.5% | 150 | 3.8% | 41 | 1.6% | 599 | 7.2% |
| Unemployed (able to work) (n, %) | 2142 | 7.3% | 72 | 0.8% | 163 | 2.7% | 117 | 3.0% | 115 | 4.6% | 1675 | 20.2% |
| Unemployed (unable to work) (n, %) | 970 | 3.3% | 164 | 1.9% | 138 | 2.3% | 32 | 0.8% | 36 | 1.4% | 600 | 7.2% |
| (b) Chronic diseases/clinical risk factors |  |  |  |  |  |  |  |  |  |  |  |  |
| At least one (n, %) | 13840 | 46.4% | 3202 | 36.4% | 2652 | 43.1% | 2019 | 51.3% | 1392 | 54.9% | 4575 | 54.6% |
| Raised blood pressure (n, %) | 6621 | 22.6% | 1841 | 21.2% | 1272 | 21.4% | 816 | 20.8% | 710 | 28.4% | 1982 | 23.9% |
| Obese (n, %) | 4844 | 16.3% | 647 | 7.3% | 1035 | 16.8% | 925 | 23.5% | 874 | 34.5% | 1363 | 16.3% |
| Diabetes mellitus (n, %) | 4642 | 15.8% | 783 | 8.9% | 741 | 12.5% | 974 | 24.8% | 414 | 16.5% | 1730 | 20.8% |
| Raised cholesterol (n, %) | 3029 | 10.4% | 364 | 4.3% | 333 | 5.6% | 247 | 6.3% | 107 | 4.3% | 1978 | 23.8% |
| CVD (n, %) | 2697 | 9.2% | 587 | 6.7% | 478 | 8.0% | 123 | 3.1% | 109 | 4.4% | 1400 | 16.9% |
| Lung (n, %) | 1470 | 5.0% | 581 | 6.6% | 278 | 4.7% | 18 | 0.5% | 28 | 1.1% | 565 | 6.8% |
| Renal (n, %) | 174 | 0.6% | 86 | 1.0% | 29 | 0.5% | 20 | 0.5% | 8 | 0.3% | 31 | 0.4% |
| Cancer (n, %) | 96 | 0.3% | 16 | 0.2% | 31 | 0.5% | 2 | 0.1% | 2 | 0.1% | 45 | 0.5% |
| (c) Behavior/risk factors |  |  |  |  |  |  |  |  |  |  |  |  |
| Current smoking (n, %) | 7185 | 24.4% | 4457 | 50.8% | 1245 | 20.9% | 281 | 7.2% | 219 | 8.7% | 983 | 11.8% |
| Current drinking (n, %) | 3247 | 11.0% | 231 | 2.6% | 1128 | 19.0% | 440 | 11.2% | 2 | 0.1% | 1446 | 17.4% |
| Body mass index (kg/m<sup>2</sup>, SD) | 25.4 | 5.7 | 23.6 | 5.1 | 25.5 | 5.4 | 27.1 | 6.6 | 28.1 | 6.5 | 25.6 | 5.2 |
| Systolic BP (mmHg, SD) | 121.3 | 19.8 | 117.4 | 19.7 | 116.2 | 18.0 | 130.4 | 20.1 | 125.3 | 18.1 | 123.7 | 19.6 |
| Diastolic BP (mmHg, SD) | 76.1 | 12.2 | 73.4 | 11.8 | 73.5 | 11.1 | 83.7 | 12.1 | 82.2 | 11.7 | 75.4 | 11.4 |
| N, % | 29809 | 100% | 8807 | 29.5% | 6152 | 20.6% | 3934 | 13.2% | 2534 | 8.5% | 8382 | 28.1% |

Note: N=Sample, SD=Standard deviation, USD=United States Dollar, CVD=Cardiovascular diseases, BP=Blood pressure, mmHg=millimeters of mercury

### COVID-19: burden, knowledge and preventive behaviours

Only 1.0% of people reported the combination of fever and cough during the study period, while 4.6% reported one or more of the recognised COVID-19 symptoms (**Supplementary Table 4**). Testing rates were consistently low, with just 0.9% of people reporting having had a swab test for COVID-19, including <10% of people with suggestive symptoms. People with known chronic disease and with higher levels of education were 50% more likely to report suggestive symptoms, even after adjustment age, gender, education and income status (P<0.01, **Supplementary Table 5**).

Awareness of COVID-19 symptoms, and of the pathways to transmission, was moderate to high for the primary components (**Supplementary Table 4**). Fever and cough were recognised as COVID-19 symptoms by 90.1% and 79.5% of people respectively. However only ∼50% of people recognised sore throat or breathlessness, and <25% tiredness, myalgia or gastrointestinal disturbance, as being potential features of COVID-19. The proportions of people unaware that COVID-19 transmission is facilitated by contaminated surfaces, or by touching the face were high (45% and 33% respectively, **Supplementary Table 4**). Weaker knowledge of symptoms and transmission was more common in women, and also strongly associated with lower levels of education, lower income status and with increasing age (**Supplementary Table 5;** odds ratio for weaker knowledge: 2.2 (95%CI: 1.9-2.4) for age >60 years, and 2.7 (95%CI: 2.4-3.3) for no formal education, compared to youngest age group and highest education group respectively (both P<0.001).

Knowledge of the personal preventive measures recommended to reduce transmission of COVID-19 was high in all settings (>95% for most metrics, **Supplementary Table 4**). However, implementation of these measures in daily life was only moderate, representing both environmental and behavioural factors (**Supplementary Table 4**). Approximately 50% of people did not have access to hand sanitisers or tissues, and 70% had no access to gloves. Almost 40% of participants did not have access to clean running water in the home, and 59% did not have access to a room that could be used for self-isolation of people with known or suspected COVID-19. In addition, 10% of people reported not wearing masks or following social distancing when outside, 25% went outside for non-essential reasons, and 75% of people continued to join meetings with people from across different households. Poorer access to and implementation of preventive measures was 2.9 (95%CI: 2.6-3.1) fold more common amongst women compared to men, and also more common amongst people from lower educational and financial backgrounds (all P<0.001; **Supplementary Table 5**).

### Impact of COVID-19 on chronic disease

Access to healthcare, including consultations, diagnostic tests and medications was compromised in all settings during the implementation of control measures (**Figure 1**). Approximately 24% of people were taking one or more regular medication, amongst whom 15% reported running out of supplies. Drugs for diabetes and hypertension were most commonly affected (61% and 44% respectively). Restrictions to movement and financial pressures were identified as the primary reasons for impaired access to healthcare and medications (**Supplementary Table 6**). The impact on access to medication was greatest in the young (OR 1.62 [95%CI:1.17-2.64] for age 18-29 years, vs age >60 years, P<0.001), in those with lower levels of income (OR 1.41 [95%CI: 1.14-1.84] for bottom vs top quintile of income, P<0.001), and education (OR 1.48 [95%CI: 1.12-2.19] for no vs 13+ years education, P<0.001), and 1.54 (95%CI: 1.12-1.95) higher amongst those with chronic disease (**Supplementary Table 5**).

**Figure 1.**
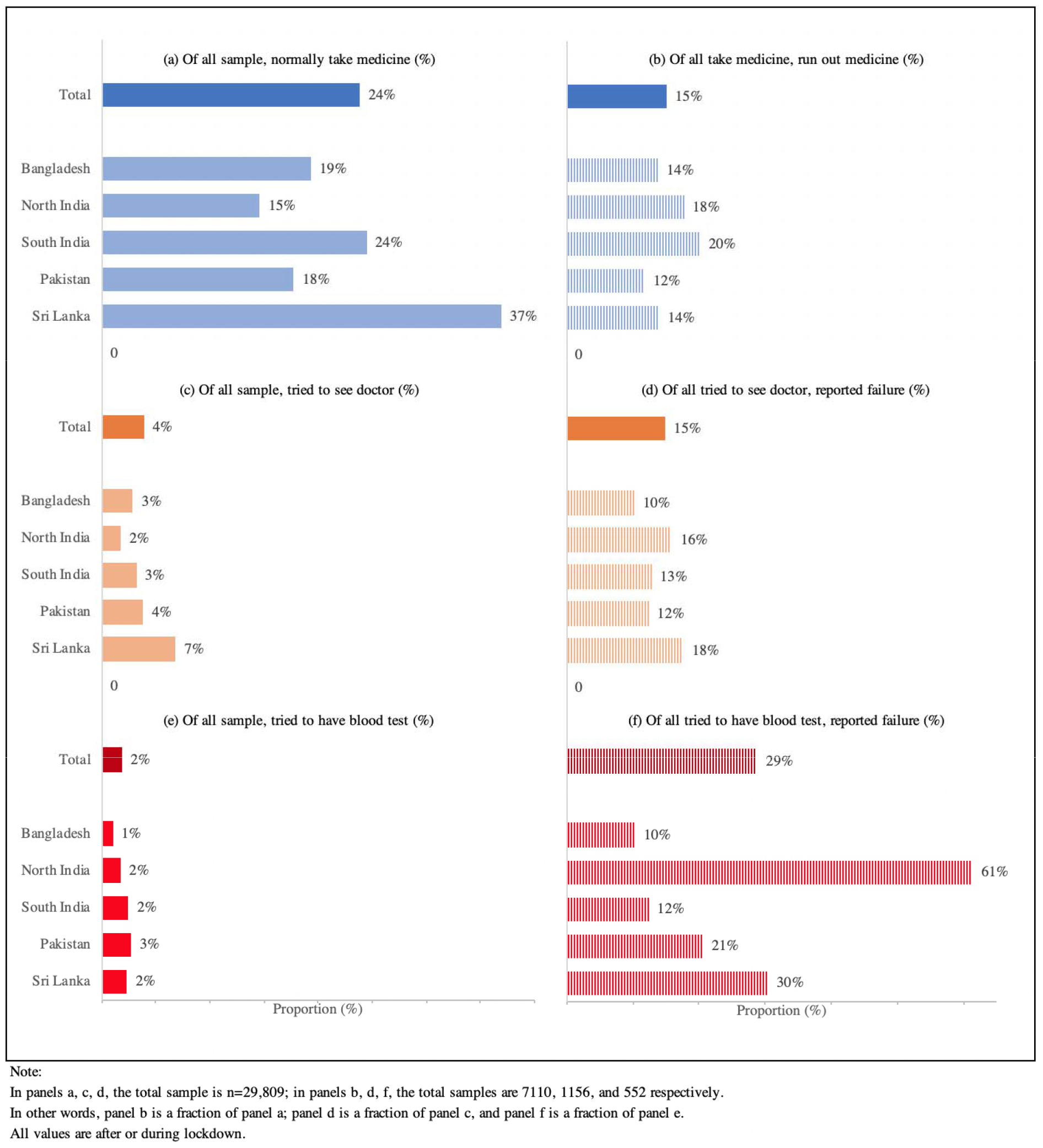
Impact of COVID-19 and control measures on access to healthcare for chronic disease.

COVID-19 also impacted behaviour patterns relevant to chronic disease (**Figure 2**). Physical activity patterns deteriorated during the pandemic, with a 32% (from 3.8 hours to 5.0 hours) increase in sedentary time, and there was a 10% (from 70% to 77.2%) increase in the proportion of people reporting inadequate fruit and vegetable intake (both P<0.001). Respondents reported weight loss in all study regions (average 3% from 62.5 kg to 62.1 kg, P<0.001). The deterioration in diet and physical activity was most marked in men (**Supplementary Table 5**). Lower levels of education were also associated with a greater negative impact of pandemic on physical activity, but a lesser effect on diet (P<0.001). Self-reported alcohol intake fell by 80% (from 11.0% to 2.2%), and smoking rates by 41% (from 24.4% to 14.5%) compared with pre-pandemic levels (both P<0.001). Continued tobacco and alcohol consumption were more than five-fold higher in men than women, while continued alcohol consumption was 1.7 fold (95%CI: 1.17-2.24) more common in the most affluent quintile compared to the lowest quintile of income (both P<0.001). If the reduction in cigarette smoking can be maintained long term, this might reduce the future risk of CVD in the population by 13%, and of lung cancer by 25% (**Supplementary Table 7**). Almost 40% of participants reported symptoms consistent with anxiety or depression (**Figure 3**). The prevalence of anxiety and depression was higher in people aged 30-49 years, amongst people with chronic disease, and in people with lower education and income (all P<0.05, **Supplementary Table 5**).

**Figure 2.**
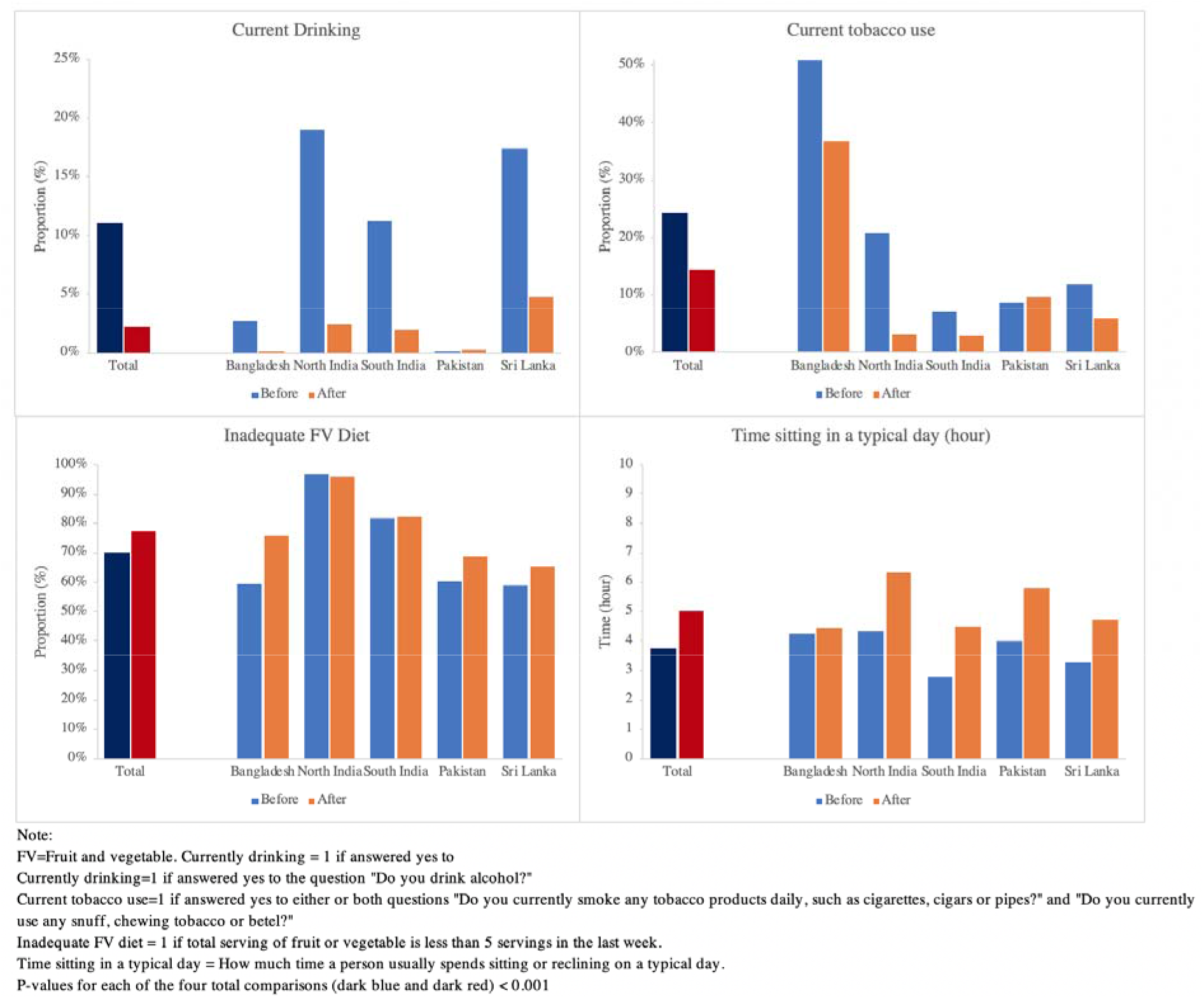
Impact of COVID-19 and control measures on behaviours relevant to chronic disease.

**Figure 3.**
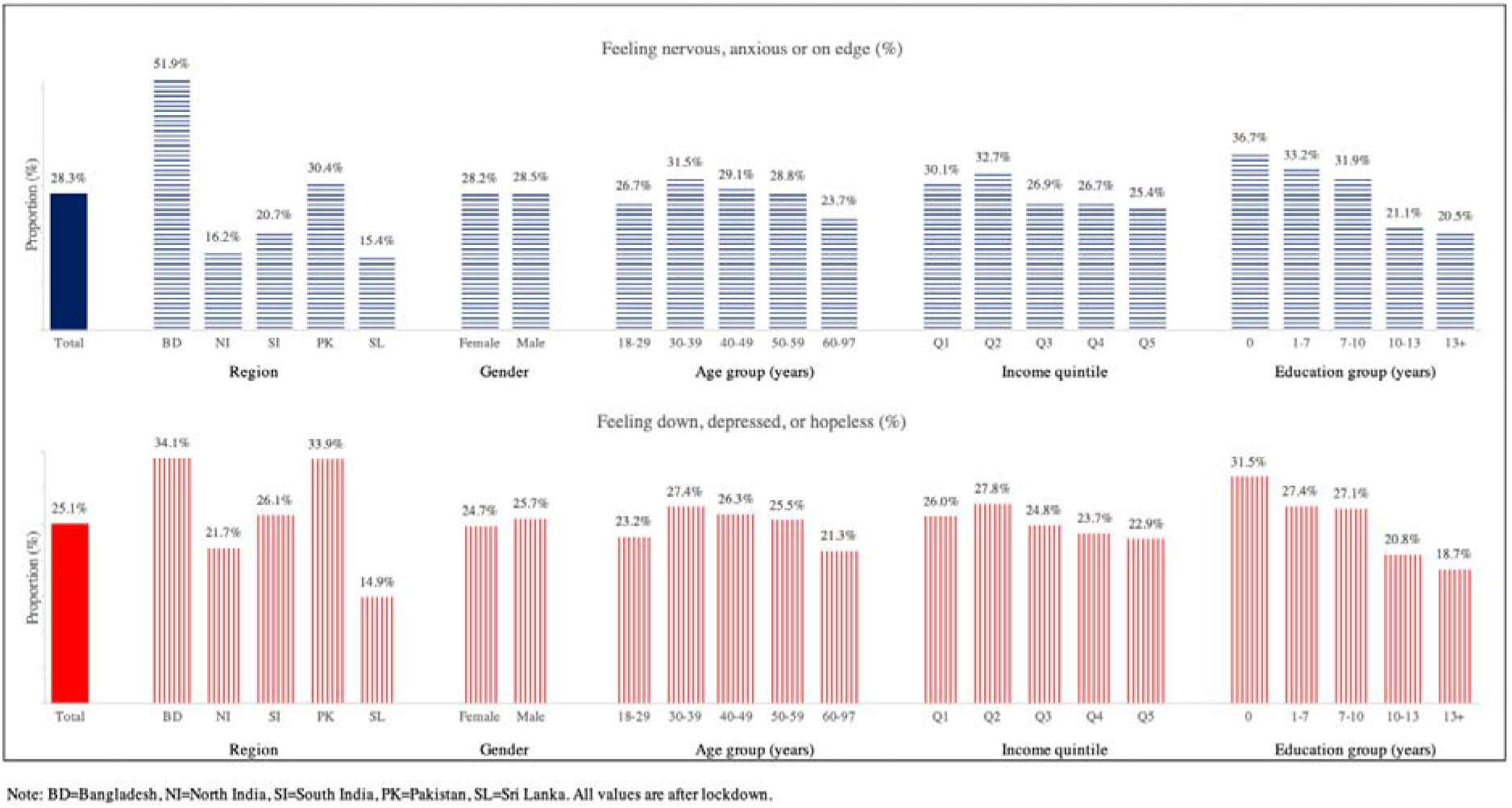
Measures of mental health during the COVID-19 pandemic.

### Impact of COVID-19 on social and financial circumstances

The impact of COVID-19 control measures on employment and incomes was high. Compared to pre-pandemic, the proportion of respondents unemployed increased 3.5 fold from 9.3% to 39.4% (P<0.001). Time in work decreased by 29% (from 41.1 to 29.2 hours) while household monthly income fell by 52% (from USD 240.0 to 114.4, P<0.001, **Figure 4**). The negative economic impact was greatest among younger participants, and amongst people with lower income and educational status at baseline (**Supplementary Table 5**). People with no formal education were 1.7 fold (95%CI: 1.53-1.97) more likely to report loss of employment, and 3.2 fold (95%CI: 2.82-3.63) more likely to report a fall in income, compared to the well-educated (both P<0.001). Almost 4% of participants had to relocate during the lockdown; of these over 80% travelled over 100 miles to their new location. Social and financial support were heterogeneous across the regions (**Supplementary Table 3**). More than 50% of people in Sri Lanka reported receiving financial support (**Supplementary Table 8**). Support with supply of medications increased 45%, whilst delivery of grocery shopping almost tripled, and access to subsidized food increased more than 6-fold. Sri Lanka also had the lowest proportion of people reporting anxiety and depression. In other settings there was little evidence of participants receiving support from state or community sources (**Supplementary Table 8**).

**Figure 4.**
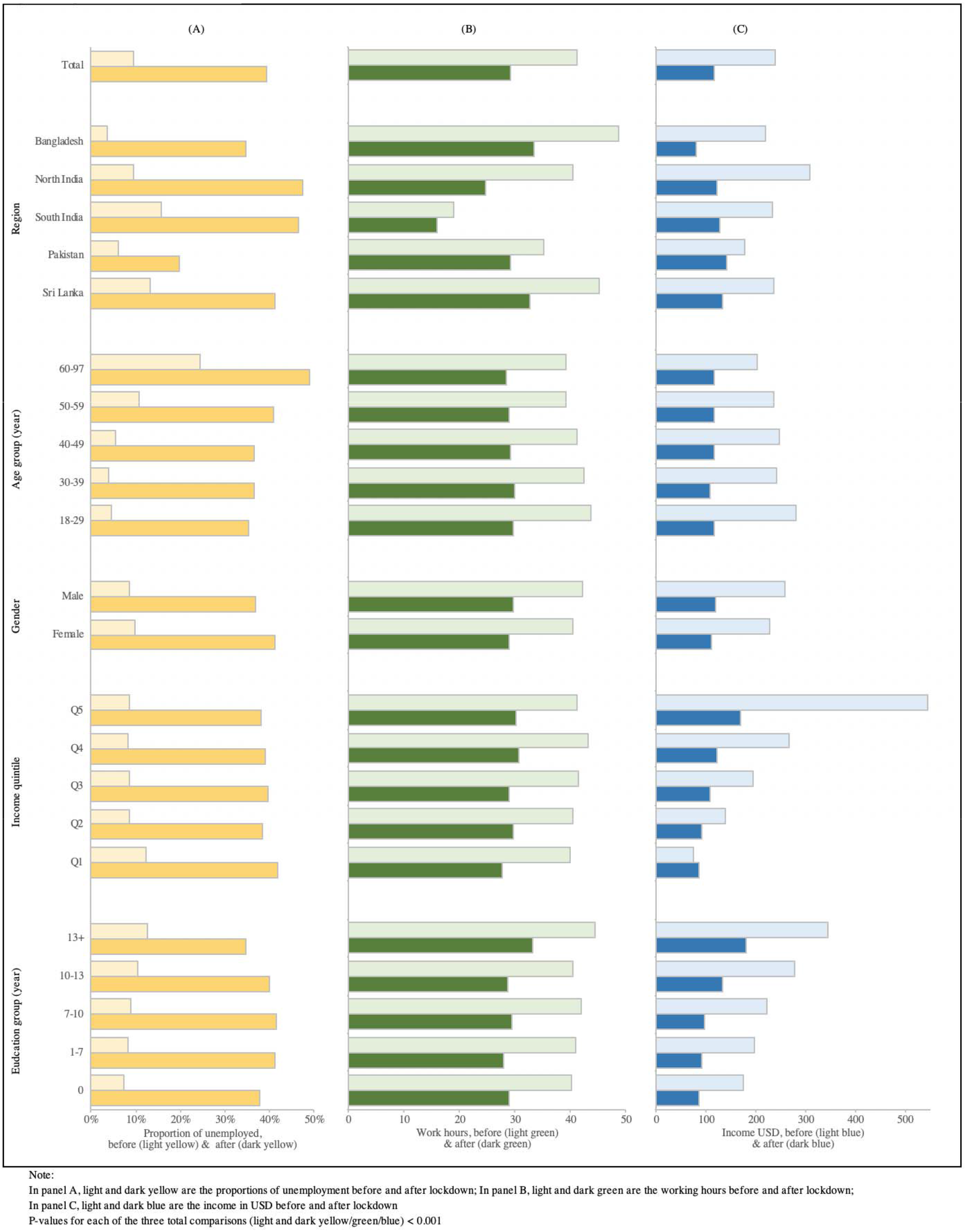
Impact of COVID-19 and control measures on employment and financial wellbeing.

## Discussion

We assessed the knowledge, behaviours, health and socio-economic circumstances of 29,809 men and women, from 93 surveillance sites in four South Asian countries, during the national lockdowns that were implemented for COVID-19 between March and July 2020. Our findings provide a comprehensive assessment of the impact of COVID-19 and pandemic response measures on the health and wellbeing of South Asian communities. We found that many South Asians had poor access to essential resources for personal protection, and that uptake of recommended preventive measures was low, especially among people from modest educational and socio-economic backgrounds. We also found major negative impacts of COVID-19, and pandemic response measures, on chronic diseases, mental health and household finances. Access to healthcare for chronic diseases was compromised, including consultations, diagnostic tests and medications. Employment, income, and working hours fell substantially in all settings, and these were accompanied by a high prevalence of anxiety and depression. Younger people, and people from lower socio-economic groups were impacted the most. Although tobacco and alcohol consumption fell, so too did physical activity levels and fruit and vegetable intake. Our findings identify factors likely to have fuelled the continued progression of COVID-19 in South Asia. They also highlight the unanticipated, inequitable and unsustainable impacts of COVID-19, and pandemic responses, on chronic disease management, mental health and socio-economic circumstances. Our findings can inform the design of future policies aimed at preventing further spread of COVID-19 in South Asia, and mitigating adverse socio-economic impacts, especially on vulnerable population groups.

### COVID-19: burden, knowledge and preventive behaviours

The prevalence of COVID-19 symptoms in the study population was low. Our results are consistent with an assessment of flu-like symptoms amongst healthcare workers in India during the same period,[12] but contrast with the findings of serology studies in Delhi and other urban South Asian settings, which indicate a high proportion of the populations tested may have been infected with COVID-19 during the lockdown.[13,14] These observations suggest that a high proportion of people in South Asia may be infected asymptomatically, in keeping with the younger age distribution of South Asian populations, compared to European or North American countries. A high proportion of individuals with asymptomatic COVID-19 represents an additional barrier to the identification and isolation of cases, underscoring the importance of molecular diagnostic assays. We note that COVID-19 testing was low during the assessment period. Although testing capacity has subsequently increased, there remain wide variations in availability, cost and uptake.

We found high knowledge for typical COVID-19 symptoms and transmission routes, positive attitudes towards preventive measures, and high adoption of hand washing with soap and water, consistent with previous reports.[15–17] We found incomplete knowledge for atypical COVID-19 symptoms and transmission routes, and important failures in the uptake of protective measures, in particular avoiding interactions between households and non-essential out-of-home activities. We found that uptake of protective measures is inversely related to education and socio-economic status, and also poorer amongst women and older people. A high proportion of respondents reported their households do not have access to running water, or suitable spaces for self-isolation. Our observations can contribute to explaining a continued sustained spread of the COVID-19 epidemic in some South Asian communities, and highlight the population groups that may benefit the most from further awareness raising measures and improved access to personal protection resources.

### COVID-19, non-communicable disease and healthy behaviours in South Asia

Almost half of participants reported at least one non-communicable disease, most commonly diabetes or hypertension, conditions known to increase morbidity and mortality from COVID-19.[18,19] In keeping with this, COVID-19 symptoms were more common amongst South Asians reporting a chronic condition. The well documented high burden of diabetes and hypertension in South Asians is contributing to the high impact of COVID-19 in this population.[20,21]

Our study design, which includes an assessment of key social and health metrics both prior to and during the pandemic, enables us to quantify the impact of COVID-19 on healthcare in different population sub-groups. We found adverse impacts on routine clinical care for people with chronic disease, reflecting a combination of reduced mobility, impaired supply of services, weakened financial circumstances and avoidance of healthcare settings. The impact was greatest amongst the more vulnerable in society, in particular those from lower socio-economic backgrounds. Our results expand the evidence base on the impacts of COVID-19 on access to healthcare in South Asia,[22–25] making a strong case for measures to protect routine health services, increase the use of digital platforms and provide medication support.[26]

Our baseline and follow-up data also enable an accurate assessment of the impact of the COVID-19 pandemic on health behaviours. We document a modest increase in inadequate fruit and vegetable consumption, and a more substantial increase in physical inactivity. Previous reports had shown increased carbohydrate consumption and snacking and reduced physical activity amongst Indians with type-2 diabetes during the pandemic.[27] However, there were also steep declines in smoking and alcohol consumption in most settings. If sustained, these might translate into a substantial reduction in the risk of chronic diseases such as cardiovascular disease and lung cancer in the population. Understanding what drove the drop in tobacco and alcohol consumption could support the development of new policies to maintain those improvements in the future, but insights will also be needed to prevent further deterioration of diet and physical activity patterns while pandemic response measures last, and to promote a return to healthy lifestyles as measures are released.

We identified a high prevalence of anxiety and depression symptoms during the study period, highest in Bangladesh. Symptoms of psychosocial distress were more common in women, and amongst people with lower income and education. Our findings are consistent with others from Bangladesh, India and Pakistan,[28] and emphasise the potential negative impact of the COVID-19 pandemic on mental health in South Asia, a region that has some of the highest suicide rates globally.[29]

### Impact of COVID-19 on social and financial circumstances

We found evidence of a high negative impact of the COVID-19 pandemic and response measures on social and economic circumstances in South Asia. We documented a substantial increase in unemployment, decreased work hours and a major reduction in household income. The adverse effects on financial circumstances were greatest for younger people, and those from less educated backgrounds. Governments throughout the world have identified the risks to economic wellbeing, and in many settings have implemented mitigation measures. Bangladesh re-opened the garment industry after just one month of lockdown, while Sri Lanka implemented a national financial support system and assistance with food supplies. However, these measures were insufficient to mitigate the tremendous impact of COVID-19 control measures on economic wellbeing. Our findings provide evidence of the scale of such impact for South Asian communities, and a strong case for robust social, organisational and fiscal measures to avoid long-lasting negative effects on the livelihoods of South Asian people.

### Strengths and limitations

We have assessed the impact of COVID-19 on behaviours, health, and wellbeing in a large representative sample of South Asians from 93 sites across four countries. This brings precision and generalisability to our findings. We benefited from the existence of comprehensive data from participants collected just before the onset of the pandemic, enabling us to accurately assess people’s changing situation. We used internationally validated questionnaires to ensure comparability with other studies. Although we recognise that the use of telephone surveys may introduce bias in recruitment, the characteristics of responders and non-responders were similar. Telephone surveys may also lead to response bias, however reliable baseline data collected pre-pandemic provided opportunities for validation in several instances. Our study was carried out at the height of the implementation of control measures, and restrictions have been eased in some countries thereafter. However, our results provide objective evidence of the impact of control measures on the population, and can inform the design and implementation of further local or national restrictions, for COVID-19 or emerging viral pandemics.

### Conclusions

Our study provides a comprehensive assessment of South Asian communities during the COVID-19 pandemic. We found a low uptake of recommended preventive measures among people from lower educational and socio-economic backgrounds, in women and in older age groups, and poor access to the resources needed for personal protection. We also found negative impacts of the pandemic on healthcare for chronic diseases, on diet and physical activity, employment and personal finances, and mental health. Impacts have been unequal, with younger people and people from lower socio-economic backgrounds impacted the most. Our results can contribute to explaining the continued progression of COVID-19 in South Asia, and provide a basis for the development of more effective, equitable and sustainable public health interventions for COVID-19 in the region.

## Data Availability

Data is available upon reasonable request.

## Acknowledgements

This research was funded by the National Institute for Health Research (NIHR) (16/136/68) using UK aid from the UK Government to support global health research, and by Wellcome Trust (212945/Z/18/Z). The views expressed in this publication are those of the author(s) and not necessarily those of the NIHR or the UK Department of Health and Social Care.

## Conflict of interest

None

## Contributions

JCC, RMA, KIK, MKM, SJ, AK, PK, FS, DK conceived the study. RP, RBK, VM, SM, SB, MH, AAMH, MS, SS, ML AM, AT, SW, SMAS, ATR, SA, CKDS, VPR, LDS, MG, BK, IYG collected and cleaned the data. DK, RP, JCC, and FS conducted data analyses. JCC, MKM, DK, and FS drafted and ML, JSK provided inputs to the manuscript. All authors approved the final version of the manuscript.

## The NIHR Global Health Research Unit for Diabetes and Cardiovascular Disease

RM Anjana, Mohan Diabetes Research Foundation, Chennai, India; John C Chambers, Imperial College London, UK; Sophie Day, Imperial College London, UK; Gary Frost, Imperial College London, UK; Sujeet Jha, Max Healthcare, Delhi, India; Prasad Katulanda, University of Colombo, Sri Lanka; Khadija I Khawaja, Services Institute of Medical Sciences, Lahore, Pakistan; Jaspal S Kooner, Imperial College London, UK; Marie Loh, Imperial College London, UK; Marisa Miraldo, Imperial College London, UK; Malay K Mridha, BRAC University, Dhaka, Bangladesh; Nicholas Oliver, Imperial College London, UK; Malibika Sarker, BRAC University, Dhaka, Bangladesh; Jonathan Valabhji, Imperial College London, UK.

---

**Supplementary Table 1:**
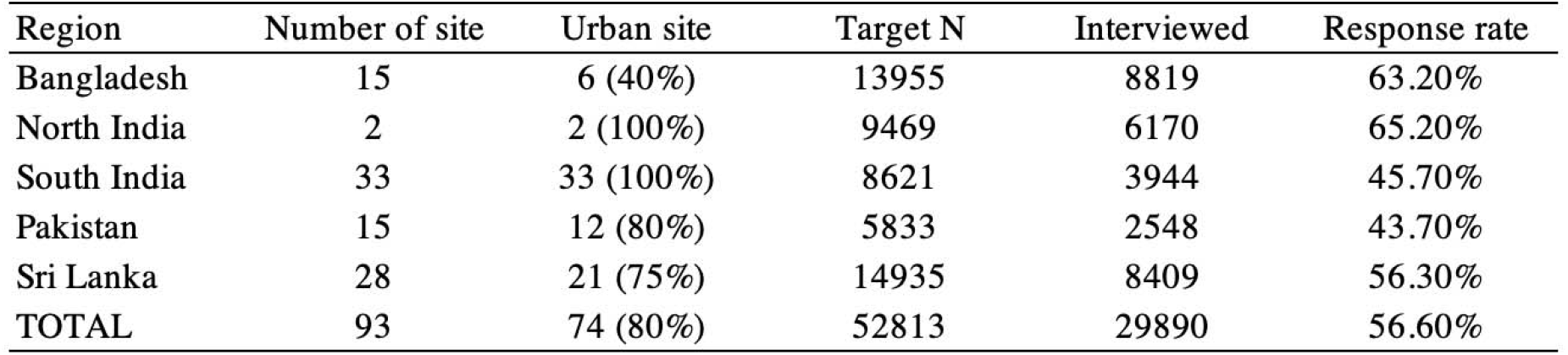
Surveilance sites by region.

| Region | Number of site | Urban site | Target N | Interviewed | Response rate |
| --- | --- | --- | --- | --- | --- |
| Bangladesh | 15 | 6 (40%) | 13955 | 8819 | 63.20% |
| North India | 2 | 2 (100%) | 9469 | 6170 | 65.20% |
| South India | 33 | 33 (100%) | 8621 | 3944 | 45.70% |
| Pakistan | 15 | 12 (80%) | 5833 | 2548 | 43.70% |
| Sri Lanka | 28 | 21 (75%) | 14935 | 8409 | 56.30% |
| TOTAL | 93 | 74 (80%) | 52813 | 29890 | 56.60% |

**Supplementary Table 2:** Characteristics of all surveilance participants (panel a) and responder bias (panel b)

|  | (a) Total baseline |  | (b) Response status |  | Crude P | Difference / OR<br>(LCI to UCI) | Adjusted P |
| --- | --- | --- | --- | --- | --- | --- | --- |
|  | n/mean | %/SD | Non-responder | Responder |  |  |  |
|  | [1] |  | [2] | [3] | [4] | [5]=[3/2] | [6] |
| N | 52,813 |  | 22912 | 29864 |  |  |  |
| Female | 33,082 | 62.6% | 65.20% | 60.80% | <0.001 | 0.84 (0.81 to 0.87) | <0.001 |
| Age (years) | 46.0 | 14.3 | 46.6 (14.5) | 45.1 (14.1) | <0.001 | -1.54 (-1.8 to -1.28) | <0.001 |
| Age categories |  |  |  |  |  |  |  |
| <30 yrs old | 7,055 | 13.4% | 13.90% | 16.70% | <0.001 | 1.15 (1.10 to 1.21) | <0.001 |
| 30-50 yrs old | 24,584 | 46.6% | 45.90% | 48.40% |  | 1.10 (1.06 to 1.14) | <0.001 |
| >50 yrs old | 21,173 | 40.1% | 40.20% | 34.90% |  | 0.83 (0.80 to 0.87) | <0.001 |
| Years at school (years) | 6.9 | 4.7 | 6.4 (4.8) | 7.3 (4.7) | <0.001 | 0.48 (0.40 to 0.55) | <0.001 |
| Education achieved |  |  |  |  |  |  |  |
| Primary school or below | 21,055 | 52.2% | 53.50% | 51.30% | <0.001 | 0.82 (0.78 to 0.85) | <0.001 |
| Secondary school | 9,104 | 22.6% | 37.80% | 39.70% |  | 1.19 (1.14 to 1.24) | <0.001 |
| Postgraduate | 10,171 | 25.2% | 8.70% | 9.00% |  | 1.11 (1.03 to 1.19) | 0.007 |
| Marital Status |  |  |  |  |  |  |  |
| Never Married | 3,578 | 6.9% | 6.40% | 7.30% | <0.001 | 1.04 (0.96 to 1.13) | 0.37 |
| Currently married | 43,823 | 84.5% | 83.40% | 85.40% |  | 1.15 (1.09 to 1.21) | <0.001 |
| Widowed | 3,869 | 7.5% | 8.90% | 6.40% |  | 0.89 (0.82 to 0.96) | 0.001 |
| Separated or Divorced | 576 | 1.1% | 1.30% | 1.00% |  | 0.78 (0.66 to 0.92) | 0.003 |
| Occupation |  |  |  |  |  |  |  |
| Government employee | 1,800 | 3.5% | 3.30% | 3.60% | <0.001 | 1.19 (1.08 to 1.31) | <0.001 |
| Non-government employee | 8,218 | 15.9% | 15.30% | 16.30% |  | 1.01 (0.96 to 1.06) | 0.75 |
| Self-employed | 13,314 | 25.7% | 25.40% | 25.90% |  | 0.93 (0.89 to 0.98) | 0.003 |
| Non-paid | 111 | 0.2% | 0.20% | 0.20% |  | 1.02 (0.69 to 1.50) | 0.91 |
| Student | 1,127 | 2.2% | 1.80% | 2.50% |  | 1.26 (1.08 to 1.46) | 0.003 |
| Homemaker | 19,744 | 38.1% | 39.30% | 37.20% |  | 1.03 (0.98 to 1.08) | 0.27 |
| Retired | 1,909 | 3.7% | 3.50% | 3.80% |  | 1.30 (1.18 to 1.44) | <0.001 |
| Unemployed (able to work) | 3,738 | 7.2% | 7.10% | 7.30% |  | 1.07 (1.00 to 1.15) | 0.06 |
| Unemployed (unable to work) | 1,890 | 3.7% | 4.10% | 3.30% |  | 0.93 (0.84 to 1.03) | 0.15 |
| Hypertension history | 11,521.0 | 22.3% | 22.60% | 22.00% | 0.13 | 1.11 (1.06 to 1.16) | <0.001 |
| Diabetes history | 8,224.0 | 15.9% | 16.40% | 15.50% | 0.004 | 1.13 (1.08 to 1.19) | <0.001 |
| Heart attack history | 1,397.0 | 2.7% | 2.80% | 2.60% | 0.37 | 1.01 (0.90 to 1.12) | 0.93 |
| Body mass index (kg/m2) | 25.4 | 5.8 | 25.4 (6.0) | 25.4 (5.7) | 0.64 | 0.46 (0.36 to 0.56) | <0.001 |
| Waist (cm) | 85.7 | 13.0 | 85.7 (13.5) | 85.6 (12.6) | 0.34 | 1.05 (0.84 to 1.27) | <0.001 |
| Systolic BP (mmHg) | 130.9 | 1961.6 | 123.5 (21.0) | 121.3 (19.9) | <0.001 | -0.02 (-0.35 to 0.32) | 0.91 |
| Diastolic BP (mmHg) | 76.6 | 12.4 | 77.2 (12.7) | 76.1 (12.2) | <0.001 | 0.38 (0.17 to 0.59) | <0.001 |
| Heart rate (bpm) | 79.0 | 13.0 | 79.4 (13.1) | 78.8 (12.2) | <0.001 | 0.25 (0.03 to 0.47) | 0.02 |
| Glucose (mg/dl) | 106.8 | 40.0 | 108.0 (41.4) | 105.9 (38.9) | <0.001 | 0.55 (-0.17 to 1.28) | 0.14 |
| Current smoker | 5,430.0 | 10.5% | 9.40% | 11.30% | <0.001 | 0.92 (0.86 to 0.98) | 0.01 |
| DrinkAlcohol |  |  |  |  |  |  |  |
| Current | 5,418.0 | 10.5% | 9.70% | 11.00% | <0.001 | 1.07 (1.00 to 1.14) | 0.07 |
| Previous | 2,143.0 | 4.1% | 3.40% | 4.70% |  | 1.15 (1.04 to 1.26) | 0.004 |
| Never | 44,290.0 | 85.4% | 86.90% | 84.30% |  | 1.09 (1.03 to 1.16) | 0.003 |
| Fruit intake (Days/week) | 3.0 | 2.5 | 2.99 (2.49) | 3.04 (2.48) | 0.03 | 0.10 (0.06 to 0.14) | <0.001 |
| Vegetable intake (Days/week) | 6.0 | 1.5 | 5.95 (1.55) | 6.06 (1.51) | <0.001 | 0.00 (-0.03 to 0.03) | 0.87 |

**Supplementary Table 3:** Control measures by region.

| Control measures in place | Bangladesh | North India | South India | Pakistan | Sri Lanka |
| --- | --- | --- | --- | --- | --- |
| Date start | 26 March 2020 | 25 March 2020 | 25 March 2020 | 16 March 2020 | 20 Mar 2020 |
| Date ended | 31 May 2020 | 31 May phased out with unlock #1 from 1 June, unlock #2 from 1 July, unlock #3 from 1 August, unlock #4 from 1 September with restrictions in containment zones till 30 September. | 31 August 2020 | 10 August 2020 | Phased out from 11 May 2020; Completed by 28 June 2020 |
| School Closing | Extended nationwide domestic restrictions on 4 August until at least 31 August amid a rise in cases, maintaining the closure of all educational institutions.<br>Confirmed on 23 August that all educational institutes will remain closed past September until the COVID-19 situation improves. | Extended closures of schools, colleges, and coaching institutions on 01 July until 31 July in both containment and non-containment zones.<br>Entered its "Unlock 3" phase of lockdown rollback on 1 August, extending nationwide closures of all schools, colleges, and educational and coaching institutions until 31 August. |  | Extended school closures on 02 July until the first week of September, past the expected reopening date of 16 July.<br>Maintained closures on educational institutes in Khyber Pakhtunkhwa and Sindh until at least 15 September despite the continued national reopening. | Began a phased reopening of schools on 06 July; following a spike in cases, schools were closed again on 13 July until at least 17 July.<br>Resumed teaching for preschool, Grade 1, and Grade 2 classes on 10 August, completing its phased national school reopening process.<br>Reopened universities on 17 August for all years |
| Workplace closing | Yes, with RMG factories started gradual opening from April 27.<br>Extended nationwide domestic restrictions on 4 August until at least 31 August amid a rise in cases, restricting business hours to 20:00 daily; closures of non-essential businesses continue to apply in designated localities under the previous colour-coded zonal lockdown system. | Allowed businesses in non-containment zones to reopen on 01 July, with official recommendations for work-from-home and the staggering of working hours; only essential services continue to be operational in containment zones.<br>Entered its "Unlock 3" phase of lockdown rollback on 1 August, permitting most businesses to reopen in all non-containment zones; recreational facilities (including cinema halls, swimming pools, bars, and assembly halls) remain closed nationwide until 31 August, while yoga institutes and gymnasiums reopened on 5 August. |  | Pakistan extended 'smart lockdowns' on 02 July for localities in some provinces, including Punjab, Khyber Pakhtunkhwa, and Sindh, mandating the closure of non-essential businesses and defining standard operating practices for essential businesses.<br>Lifted the complete lockdown in Punjab province on 3 August; imposed in advance of the Eid-al-Adha holiday, the lockdown was rolled back two days earlier than scheduled, allowing all businesses to reopen weekdays from 9:00 to 19:00 (except for recreational facilities, including restaurants, beauty parlours, and cinemas).<br>Lifted restrictions on restaurants and some non-essential tourism-related businesses in the week following 10 August, while Khyber Pakhtunkhwa, Punjab, and Sindh provinces maintained some restrictions on marriage halls and expo centres until 15 September. | Yes |
| Cancel Public Events | Extended nationwide domestic restrictions on 4 August until at least 31 August amid a rise in cases, maintaining a ban on all large public gatherings. | Announced the cancellation of all social, political, sports, religious, entertainment, academic and cultural events from 01 July in both containment and non-containment zones<br>Entered its "Unlock 3" phase of lockdown rollback on 1 August, extending its nationwide prohibition of public events (including social, political, entertainment, cultural, and religious functions) until 31 August. |  | Announced new requirements on religious congregations in Khyber Pakhtunkhwa, Punjab, and Sindh, with all festivals permitted with the condition of prior approval from respective provincial governments and adherence to local SOPs |  |
| Restrictions on Gatherings | Extended nationwide domestic restrictions on 4 August until at least 31 August amid a rise in cases, maintaining a ban on large private gatherings; restrictions of private gatherings to fewer than 10 people continue to apply in designated localities under the previous colour-coded zonal lockdown system. | Extended a ban on large public gatherings on 01 July, while restrictions on weddings were loosened to no more than 50 guests, and funerals to no more than 20 guests<br>Maintained its restrictions on private gatherings under its National Directives for COVID-19 Management on 1 August, including limits on marriage-related gatherings at 50 people, and funerals at 20 people. |  | Announced on 12 August a phased approach to lifting sports-related gatherings as part of its lockdown reopening; in the current first phase, 10 or fewer people are permitted at workouts and practices nationwide |  |
| Close public transport | Resumed limited public transport services on 4 August; capacity reductions and social distancing measures are in place on all rail services, and on bus services operating in Dhaka and on long-distance routes. | Resumed domestic flights and passenger trains on 01 July, metro rail systems remain closed.<br>Entered its "Unlock 3" phase of lockdown rollback on 1 August, extending its nationwide closure of all urban "Metro Rail" services until 31 August; domestic trains and other local transports continue to adhere to regional Standard Operating Procedures (SOPs) issued by the Ministry of Health and Family Welfare. |  | Continued reopening all public transport since 10 August with SOPs for mitigating COVID-19 transmission in place |  |
| Stay at home requirements | BANGLADESH: extended nationwide domestic restrictions on 4 August until at least 31 August amid a rise in cases, recommending all residents to stay home "to the extent possible"; requirements to stay at home continue to apply in designated localities under the previous colour-coded zonal lockdown system. | Imposed a nightly curfew nationwide on 01 July, and general daytime movements were restricted in containment zones except for the operation of essential services.<br>Entered its "Unlock 3" phase of lockdown rollback on 1 August, extending mandatory stay-at-home requirements in all containment zones nationwide until 31 August, except for "essential activities". |  | Directed citizens to stay confined in homes when subject to 'smart lockdown' notifications in their locality as of 02 July.<br>Extended a complete lockdown in Balochistan province on 3 August until 17 August, mandating residents to stay at home; all recreational places (including beaches) reopened in Sindh province on 4 August, ending a lockdown imposed in advance of the Eid-al-Adha holiday.<br>Continued its withdrawal of stay-at-home measures across various provinces since the 10 August reopening. |  |
| Restrictions on Internal movement | Yes | Maintained strict perimeter control in containment zones as of 01 July, barring movement in or out of designated areas.<br>Entered its "Unlock 3" phase of lockdown rollback on 1 August, maintaining "strict perimeter control" of all containment zones nationwide until 31 August; all movement in or out of these zones remains prohibited except for medical emergencies and the supply of essential goods and services.<br>Requested state governments to lift all remaining inter-state restrictions on the movement of goods and their conveyances in compliance with a mandate under the current "Unlock 3" lockdown rollback by the Ministry of Home Affairs |  | Formalized restrictions on internal movement in its 'smart lockdown' strategy as of 02 July, barring movement outside containment areas | Yes |
| International Travel Controls |  | Extended its ban on all international flights on 01 July with the exception of repatriation flights.<br>Entered its "Unlock 3" phase of lockdown rollback on 1 August, maintaining its ban on all international air travel of passengers nationwide, except where permitted by the Ministry of Home Affairs (including for the repatriation of citizens). |  |  |  |
| Financial Support | Yes (for low SES) | Yes (to beneficiaries of specific national schemes and people with below-poverty-line cards) | Yes (for low SES) | Yes (for low SES) | Yes (for low SES and informal workers) |
| Nutritional Support | Yes | Yes | Yes | Yes (for low SES) | Yes (for low SES and informal workers) |

Note: SES = Socioeconomic status
Source: Oxford COVID-19 Government Response Tracker: Regional Report - South Asia & Country collaborators https://www.bsg.ox.ac.uk/research/research-projects/coronavirus-government-response-tracker

**Supplementary Table 4:**
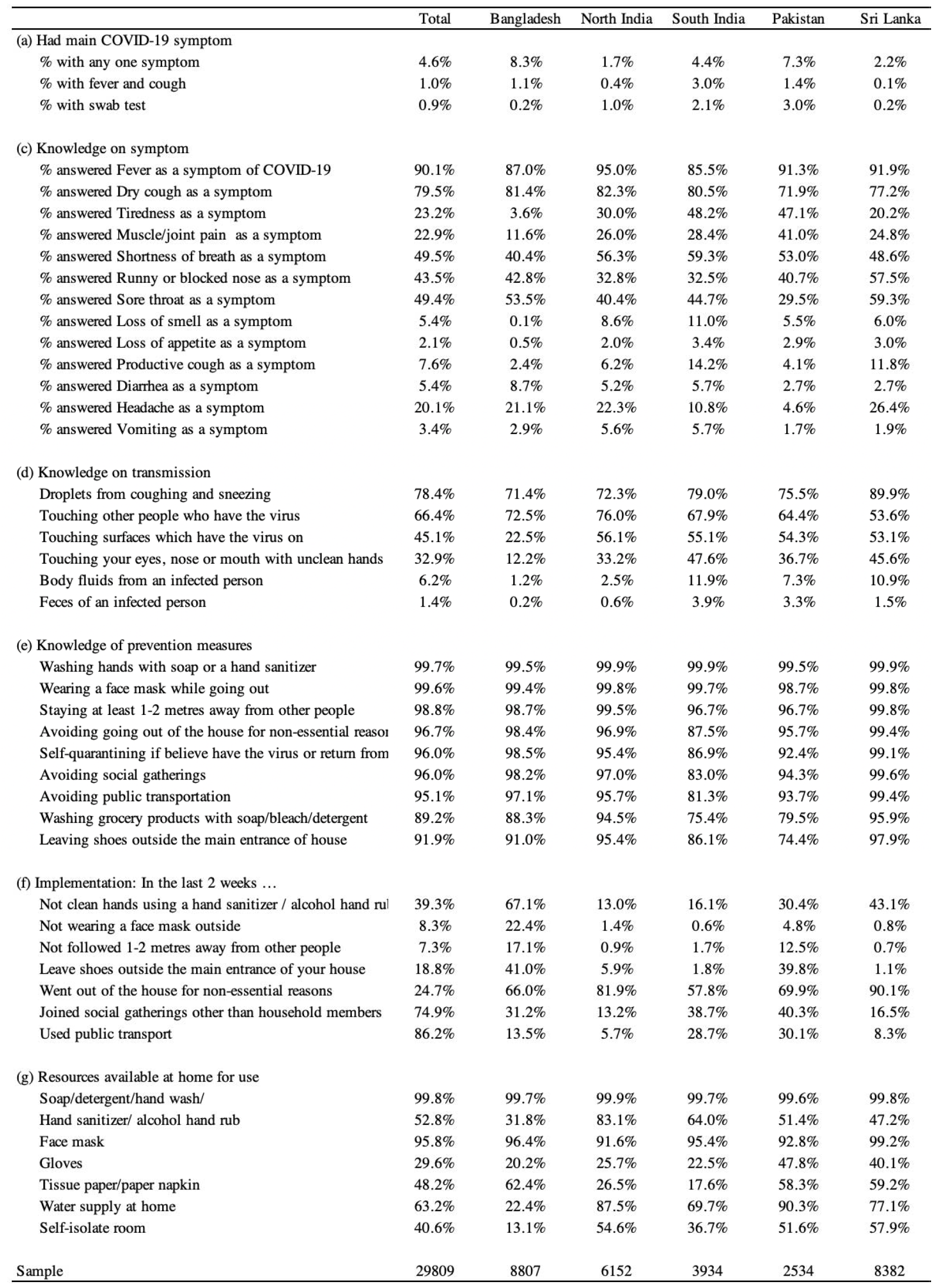
COVID-19 Burden, knowledge and preventive behaviours.

**Supplementary Table 5:**
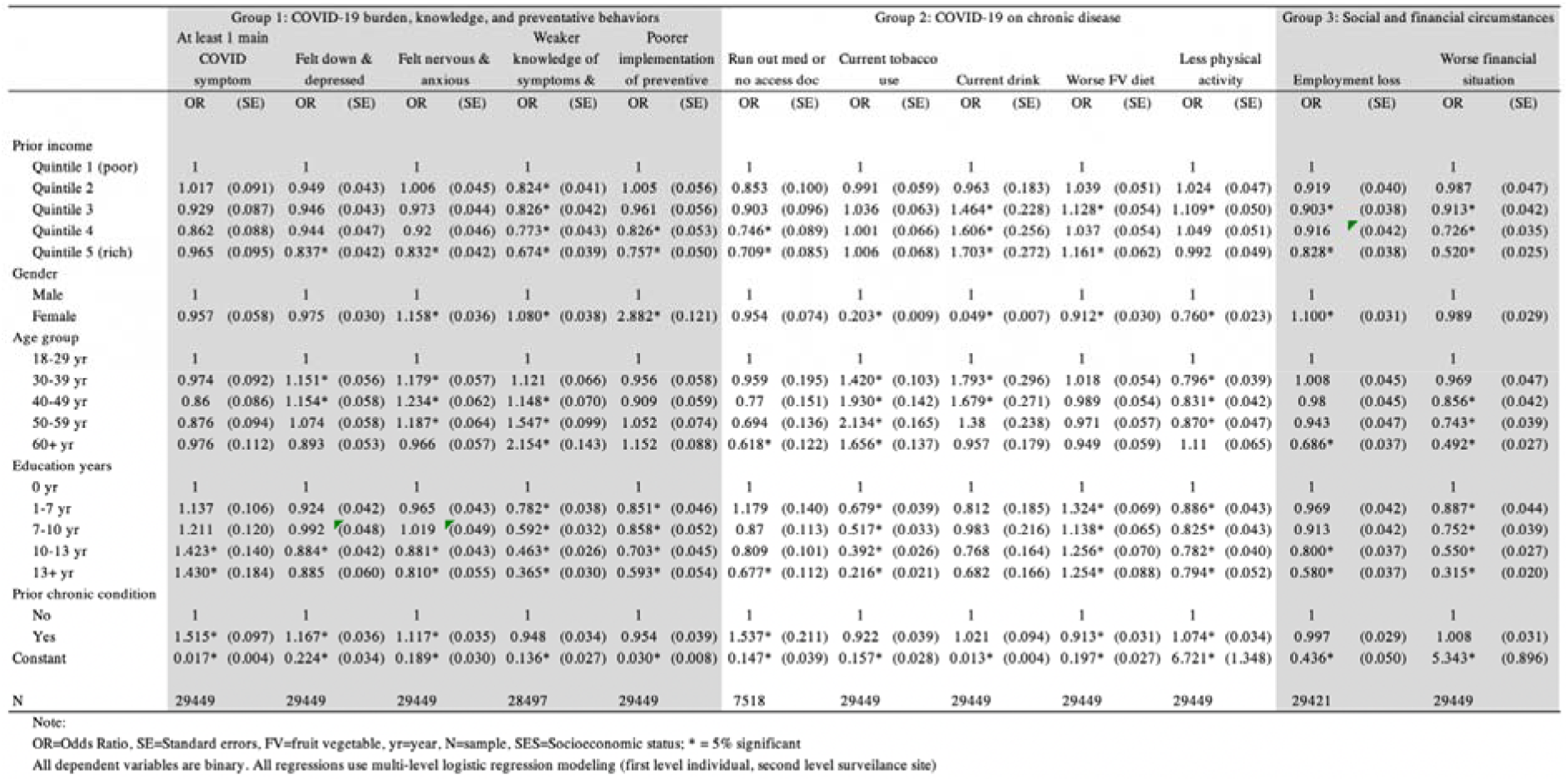
COVID-19 related lockdown impacts on soclocconomic and health status in South Asia.

**Supplementary Table 6:** COVID-19 and access to medications.

|  | Total | Bangladesh | North India | South India | Pakistan | Sri Lanka |
| --- | --- | --- | --- | --- | --- | --- |
| (a) Of all sample, on routine medications | 23.9% | 19.3% | 14.6% | 24.5% | 17.8% | 36.9% |
| (b) Of those on routine meds, run out of any meds (%) | 15.1% | 13.8% | 17.7% | 20.1% | 11.8% | 13.9% |
| (c) Of those run out of meds, types of medications |  |  |  |  |  |  |
| Run out: Anti-diabetic | 60.9% | n/a | 56.2% | 76.5% | 68.6% | 53.5% |
| Run out: Anti-hypertensive | 43.8% | n/a | 40.5% | 34.8% | 58.8% | 47.3% |
| Run out: Aspirin or clopidogrel | 8.6% | n/a | 2.5% | 10.7% | 9.8% | 9.5% |
| Run out: Statin | 8.2% | n/a | 2.5% | 0.5% | 2.0% | 14.9% |
| Run out: Pain killers | 6.7% | n/a | 9.1% | 3.7% | 11.8% | 6.8% |
| Run out: Inhaler | 5.6% | n/a | 4.1% | 1.6% | 0.0% | 8.9% |
| Run out: Cough medications | 1.6% | n/a | 3.3% | 0.0% | 2.0% | 1.9% |
| (d) Of those run out meds, reason for running out |  |  |  |  |  |  |
| Lockdown restricted me from going out | 50.9% | 8.5% | 55.4% | 71.7% | 17.0% | 67.1% |
| Unable to afford | 16.7% | 48.5% | 6.3% | 4.1% | 47.2% | 5.1% |
| Unable to go out for other reasons | 9.1% | 17.0% | 6.9% | 5.2% | 17.0% | 6.5% |
| Was afraid of going out due to fear of COVID-19 infecti | 7.7% | 3.8% | 13.8% | 2.6% | 13.2% | 9.3% |
| Pharmacy out of stock | 6.8% | 5.5% | 10.1% | 7.2% | 3.8% | 6.5% |
| Others | 4.7% | 16.2% | 1.3% | 1.6% | n/a | 1.6% |
| Government medicine out of stock | 4.1% | 0.4% | 6.3% | 7.7% | 1.9% | 3.9% |
| Sample | 29809 | 8807 | 6152 | 3934 | 2534 | 8382 |

Note: n/a: Data not available

**Supplementary Figure 7:**
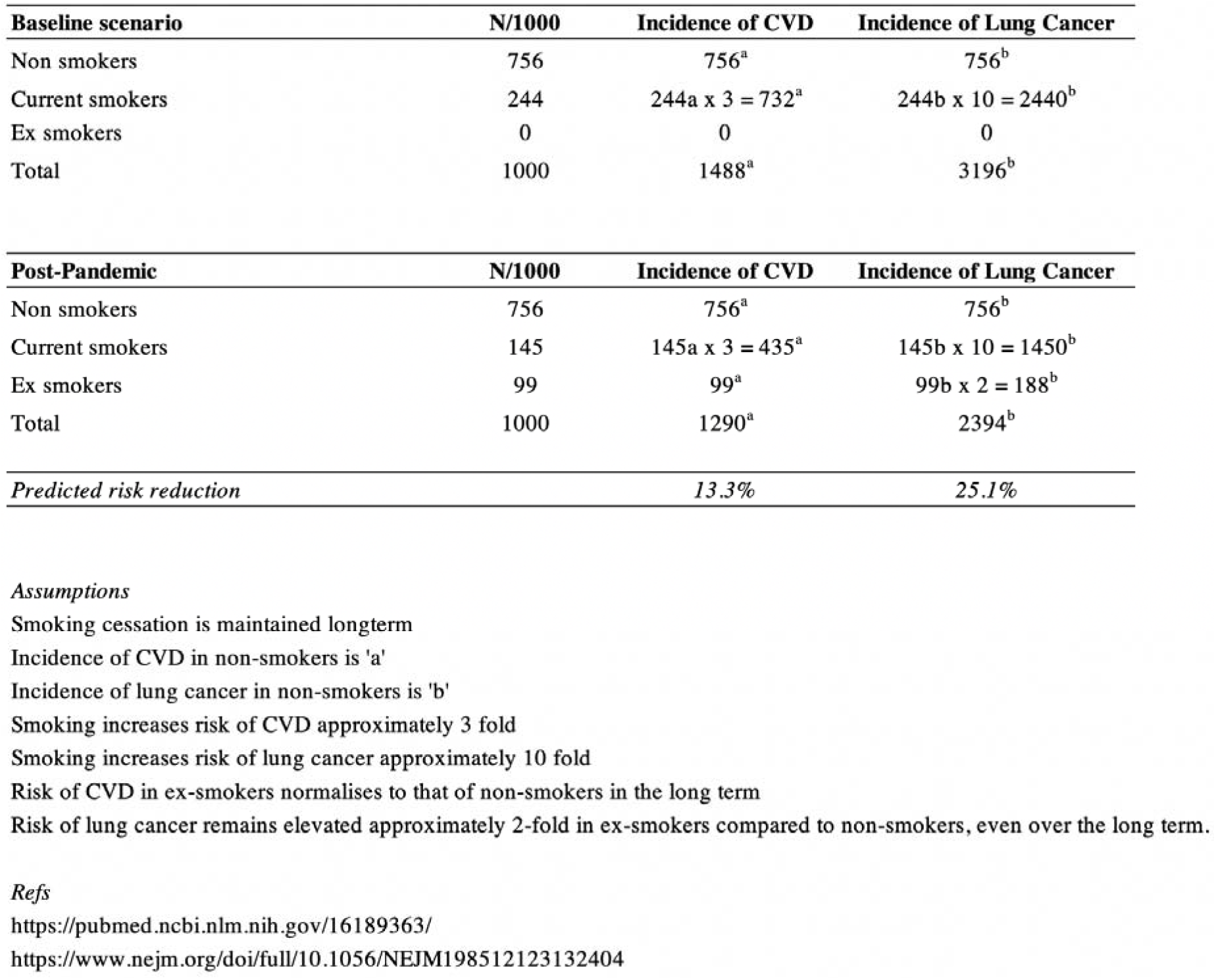
Benefit calculation of smoking cessation.

**Supplementary Table 8:** COVID-19 and social and financial support.

|  | Total | Bangladesh | North India | South India | Pakistan | Sri Lanka |
| --- | --- | --- | --- | --- | --- | --- |
| (a) Shopping for groceries and food |  |  |  |  |  |  |
| Before lockdown | 20.6% | 3.8% | 26.6% | 64.4% | 17.4% | 14.2% |
| After lockdown (Last 2 weeks) | 26.9% | 7.2% | 26.7% | 49.8% | 16.7% | 40.0% |
| % Difference | 30.7% | 91.0% | 0.3% | -22.6% | -4.1% | 181.9% |
| (b) Picking up medications |  |  |  |  |  |  |
| Before | 10.2% | 2.7% | 5.0% | 38.2% | 9.4% | 9.1% |
| After | 8.7% | 1.7% | 3.9% | 23.3% | 7.4% | 13.2% |
| % Difference | -14.6% | -35.0% | -22.3% | -38.9% | -21.1% | 44.6% |
| (c) Donated or subsidised food |  |  |  |  |  |  |
| Before | 6.1% | 3.7% | 10.1% | 12.4% | 5.2% | 3.0% |
| After | 13.2% | 7.6% | 12.6% | 14.8% | 4.2% | 21.5% |
| % Difference | 115.7% | 103.3% | 25.0% | 19.3% | -19.5% | 611.9% |
| (d) Monetary support |  |  |  |  |  |  |
| Before | 6.6% | 3.7% | 3.3% | 19.4% | 2.7% | 7.3% |
| After | 20.1% | 3.3% | 4.4% | 21.2% | 2.4% | 54.2% |
| % Difference | 203.1% | -12.2% | 33.2% | 9.4% | -11.6% | 638.2% |
| Sample | 29809 | 8807 | 6152 | 3934 | 2534 | 8382 |

Note: P-values for each % difference among total sample < 0.001

**Supplementary Figure 1:**
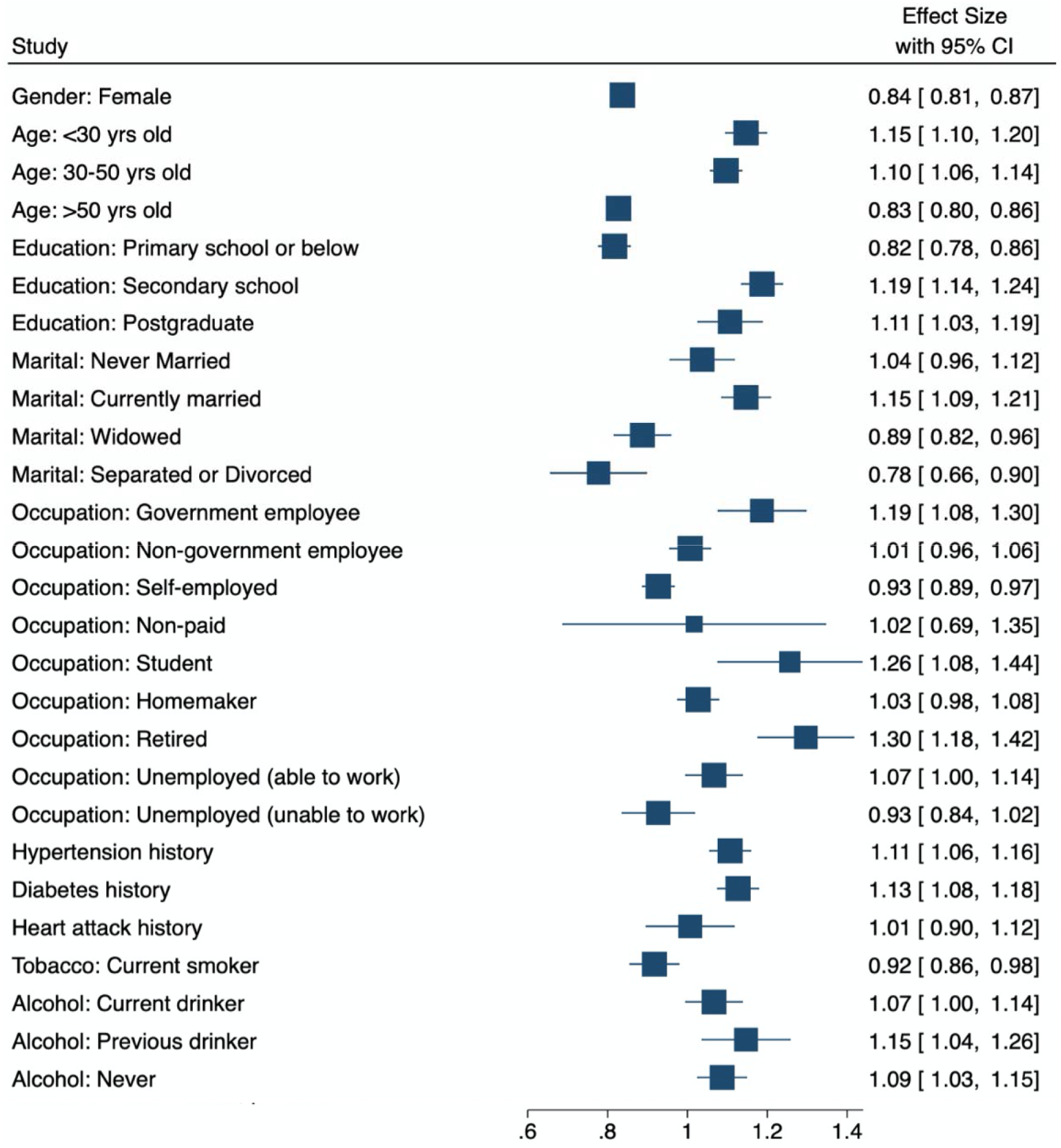
Forest plot of responser bias.

## Notes

### Competing Interest Statement

The authors have declared no competing interest.

### Author Declarations

Research approval has been obtained from the Imperial College London Research Ethics Committee (reference: 18IC4698) and local Institutional Review Boards in each of the participating countries: BRAC University IRB (ref: 2018 013 IR) Max Healthcare Institutional Ethics Committee (ref: RS/MSSH/SKT-2/ENDO/IEC/18-39 MDRF Institutional Ethics Committee (ref: IRB2640) SIMS IRB (ref: 2018/424/SIMS) Univ of Colombo Ethics Review Committee (ref: EC-18-094)

